# Did a true change occur? Improving analytical decisions for serial performance testing in sport

**DOI:** 10.1101/2025.09.22.25336149

**Authors:** Patrick Ward, Scot Morrison, Garrett Bullock

## Abstract

Monitoring strategies are used to assess athletes’ physical readiness to maximize performance. These data are used to assist practitioners and coaches in making decisions about the volume and intensity of the upcoming training, practice, and competition. Often, these measurements are serial in nature, where the athlete is evaluated for longitudinal changes over the course of a competitive season. The target of these strategies is commonly directed at determining whether the athlete has recovered from the most frequent bout of training or competition or adapted to the current training. Sport scientists seek to identify individualized athlete changes from serial measurements to understand how the athlete is responding to their training and competitive demands to optimize performance. Within this paper, we demonstrate novel individualistic probabilistic Bayesian methods that improve analytical based decisions concerning true athletic changes during serial (i.e., repeated) measures. We then discuss practical examples, providing open access computer code and data, to assist the practitioner in creating future scalable resources.

## Introduction

Professional and collegiate sports represent a high-stakes environment, where winning is critical to improving the financial vitality of the team and ensuring coaches and players retain their jobs. Consequently, the availability of the best players to be their best come game day is of the highest importance.(Ward et al., 2018) This has created an interest in monitoring strategies to assess athletes’ physical *readiness* and track progress over time in an effort to mitigate injuries or decreases in performance.(Swinton et al., 2018; Weakley et al., 2024; Weakley et al., 2021) These data are used to assist practitioners and coaches in making decisions about the volume and intensity of the upcoming practice week.(Weakley et al., 2024) Often, these measurements are serial in nature, where the athlete is evaluated for longitudinal changes over the course of a competitive season.(Datson et al., 2022) The target of these strategies is commonly directed at exploring whether the athlete has recovered from the most frequent bout of exercise or competition or adapted to the current training.(Harry et al., 2024)

McLean and colleagues,(McLean et al., 2010) used a weekly counter movement jump test to identify decrements in neuromuscular output in professional Rugby athletes at 24, 48, and 72-hours post-match. They found that Rugby athletes often exhibit declines in neuromuscular performance at 48 hours following competition, indicating that coaches and practitioners should be sensitive to the training intensity applied during this time given that the athletes are not completely recovered.(McLean et al., 2010) Similar approaches to understanding when neuromuscular output has returned to baseline following competition have been explored in Australia football,(Kinsella et al., 2012) Soccer,(Armada-Cortés et al., 2022) and Basketball.(Sanders et al., 2019) Unlike medical evaluations, where there is a defined endpoint (e.g., injury or illness),(Mchunu et al., 2022) these monitoring approaches in sport science are often devoid of a target outcome. Rather, the practitioner is observing an underlying process over time (i.e., neuromuscular recovery or strength gains) and attempting to react to changes in that process if outcomes differ from what would normally be expected.(Hecksteden, 2017; McLean et al., 2010) For example, changes in the countermovement jump test are not directly linked to in-game performance, but instead signal practitioners that deleterious changes that limit physical ability may be occurring.(McLean et al., 2010)

Ultimately, the practitioner is seeking to answer the question, *“How confident are we that the change we observe today is meaningful enough to intervene or make an adjustment to the planned training program?”* Answering this question is not an easy task, as analyzing such longitudinal data requires the sport scientist to consider statistical approaches that may be foreign to what they’ve been taught during their education. Most statistics courses in sport science university programs focus on analyzing group-level data,(Harry et al., 2024) as in well-designed studies that describe the average behavior of a sample population.(Beltz et al., 2016) Often, this leads to analysis which compares the observed change to a set threshold, such as the Smallest Worthwhile Change (SWC),(Turner et al., 2015) Coefficient of Variation,(Brown, 1998) or Typical Error of Measurement (TEM).(Perini et al., 2005) While such approaches to analysis may be useful for broader inference, sport deals with individuals and group analysis often lacks generalizability to the individual level.(Bullock et al., 2023)

The interest in individual athlete analysis and limitations of group-ba<u>s</u>ed analysis in elite sport has led to more attention being placed on single subject or N-of-1 research designs.(McDonald et al., 2020) These methods have been commonly used in medicine, social work, and psychology. Recently, Bullock and colleagues(Bullock et al., 2023) suggested the use of single subject analysis and natural experiments in elite sport environments, where randomized controlled trials would not be feasible. Additionally, individualizing such analysis allows for the utilization of probabilistic measures, to aid in decision-making.(Ward et al., 2018) Threshold-based outputs, such as comparing the change to an smallest worthwhile change (SWC) or typical error of measurement (TEM) lead to dichotomous thinking where the athlete has either observed a change or not. However, modeling the probability that the observed performance is meaningfully different from a threshold value allows for a more comprehensive discussion of risk.(Sottas et al., 2007) For example, Heckstenden et al.,(Hecksteden, 2017) have applied a Bayesian approach to assessing fatigue in elite soccer athletes, weighting prior knowledge and observed creatine kinase levels to leverage a posterior distribution that explains the probability of the observed outcome.

Sport scientists require methodological approaches that can identify individualized athlete changes from serial measurements to understand how the athlete is responding to their training and competitive demands.

There is currently a paucity of sport analytical methods that adequately assist the sports scientist in determining if a true change occurred in longitudinal training or competition. This inhibits sports scientists’ ability to make informed decisions and optimize athlete performance. Thus, in this paper we demonstrate novel individual-level probabilistic Bayesian methods that improve practical decision-making about true athletic changes across serial (i.e., repeated) measures. We then discuss practical examples, providing open access computer code and data, to assist the practitioner in creating future scalable resources.

## Materials and Methods

### Analytical Considerations

There are multiple statistical considerations when integrating serial testing at the individual level. A list of key definitions are reported in Table 1.

**Table 1.**
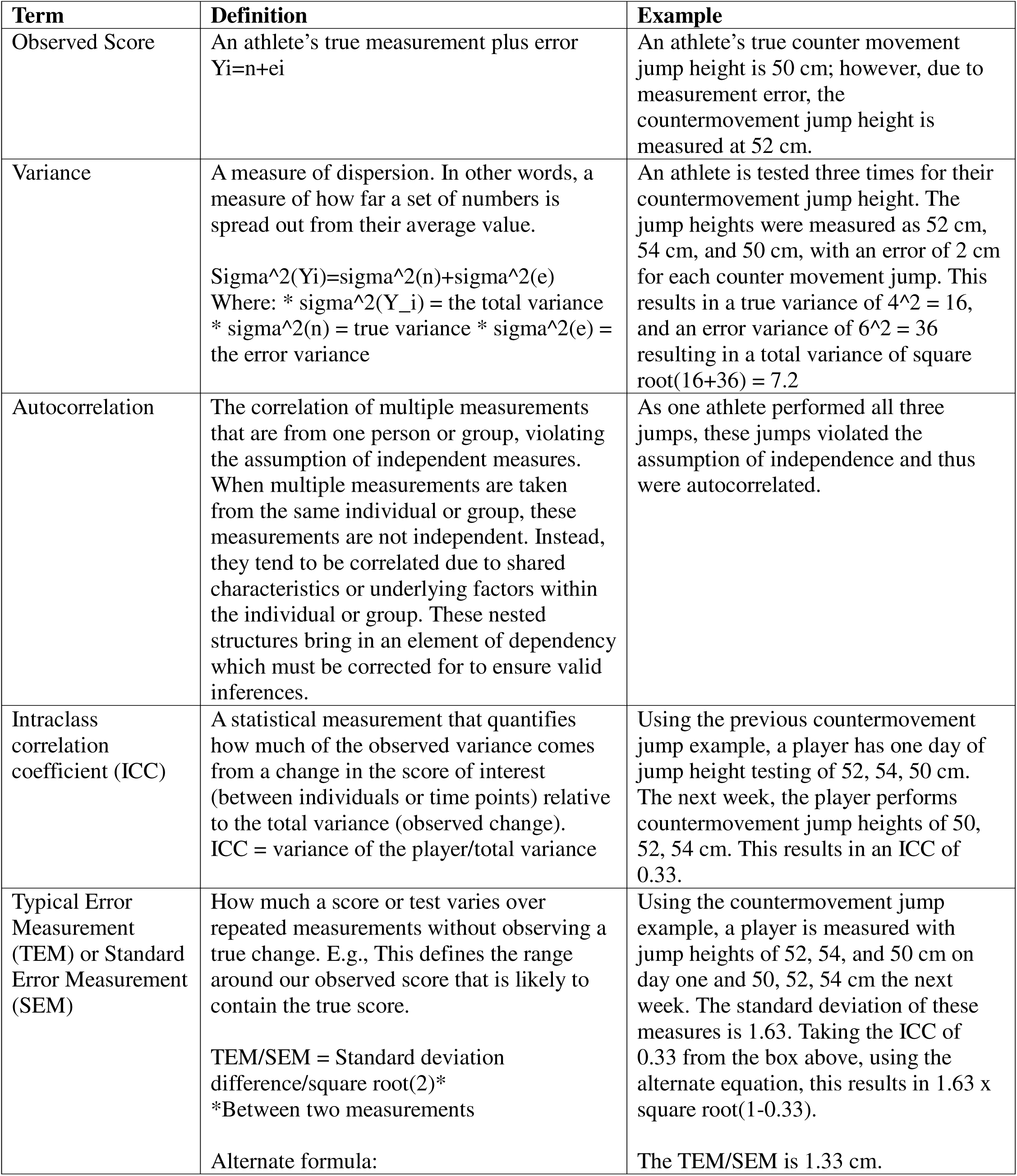
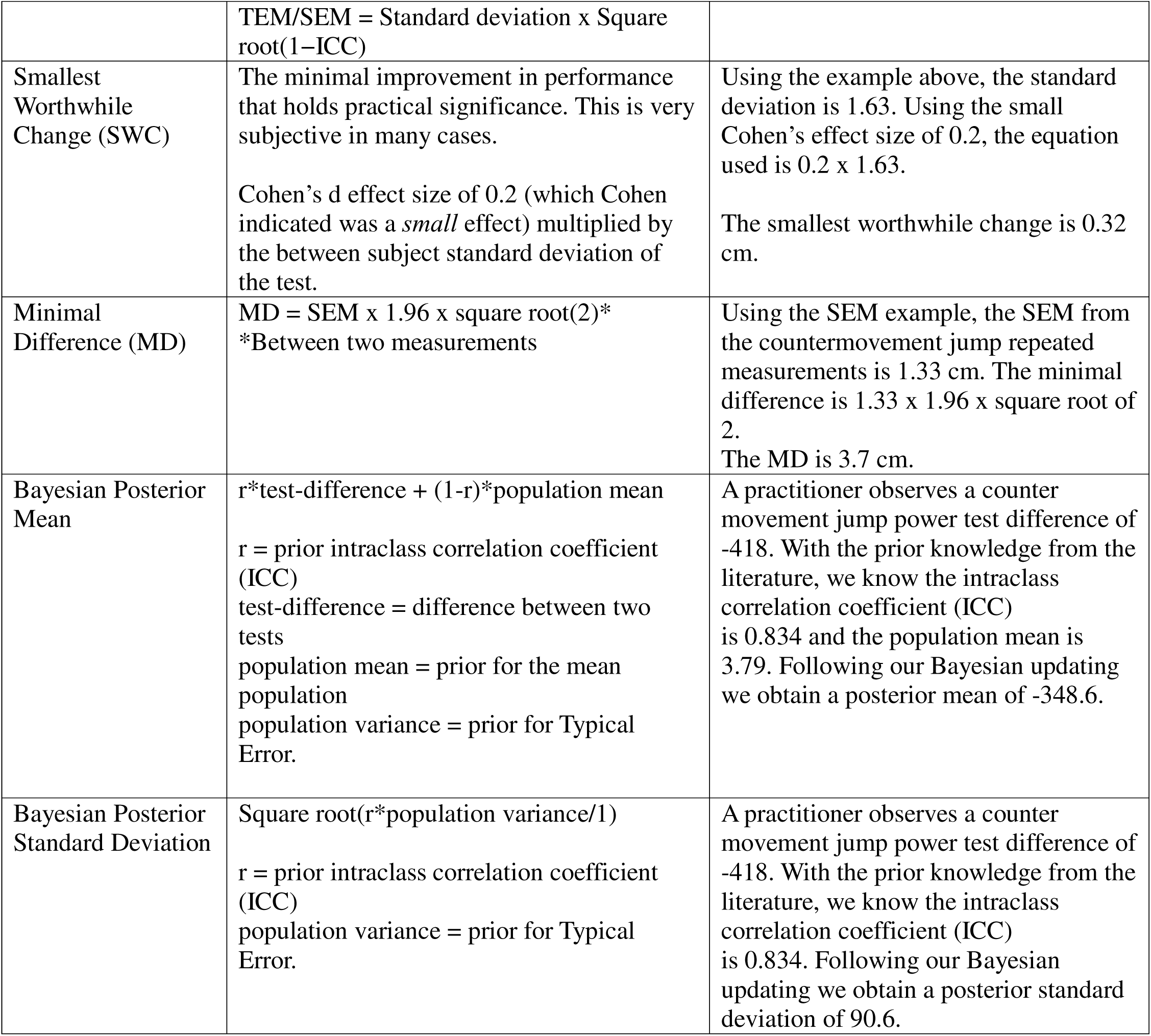
Key Analytical Definitions.

### Single Subject Analysis

Single subject analysis, also known as idiographic or more commonly as N-of-1 methods, utilize repeated measures from one participant, allowing individualized conclusions about specific tests and interventions.(Beltz et al., 2016) N-of-1 designs are different than case series, as precise conclusions, using scientific rigorous methods are employed.(Bullock et al., 2023; McDonald et al., 2020) N-of-1 approaches have previously been applied in fields such as personalized medicine(Van Der Greef et al., 2006) and psychology,(Woodworth et al., 2016) allowing for a generalizable and scalable scientific method. Moreover, such approaches are directly meaningful to the practitioner who often use published scientific findings coupled with the unique information being observed or reported by the athlete they are working with to build an individualized approach to treatment or training.(Bullock et al., 2023) Incorporating N-of-1 strategies for serial sport testing therefore has the potential to reveal individual athlete specific changes.

### Bayesian Versus Frequentist Approaches

Statistical worldviews, and the approaches that come from them, can be broadly categorized as either frequentist or Bayesian.(Bland & Altman, 1998) Both offer unique advantages and disadvantages, which we will review here.(Bland & Altman, 1998) In the most general sense, we can consider frequentist approaches to be reliant on the long run frequency to establish probabilities which are considered to be a property of the data.(Bland & Altman, 1998) In contrast, the Bayesian approach sees probabilities as representing a degree of belief (or uncertainty) that we have about an estimate given the available data and our prior knowledge.(Gelman et al., 1995)

Scientific approaches to identifying whether an outcome observed on a test is outside of a reference range are commonplace in health and medicine.(Hecksteden, 2017) Unfortunately, most of these approaches rely on a single observation relative to a population-derived threshold used to differentiate between “abnormal” and “normal” responses. Such an approach leads to dichotomous thinking, fails to account for uncertainty and error in the test procedure, and limits the practitioner’s ability to make probabilistic statements regarding their level of confidence in the recent observation.(Altman & Royston, 2006) Assessing the probability that a recent test observation is outside of normal can be useful for making decisions about potential interventions and has been discussed in both cancer research(McIntosh et al., 2024; McIntosh et al., 2025) and positive Olympic drug testing.(Sottas et al., 2007)

Given the serial nature of repeated testing and within-individual correlation, statisticians have advocated for dynamically adapting reference ranges, using Bayesian methodologies, to combine both prior knowledge of the test in the specific population as well as the individual’s repeated testing history.(Schechter & Adler, 1988) McIntosh et al. (2024, 2025)(McIntosh et al., 2024; McIntosh et al., 2025) have applied such an individualized approach to repeated cancer screening, allowing for earlier identification of cancer diagnoses compared to traditional thresholds. Similarly, Sottas et al.(Sottas et al., 2007) developed the Athlete Passport for the International Olympic Committee, enabling faster identification of athletes violating doping regulations. These methodologies have also influenced sport science research, including the probability of fatigue or recovery (Heckstenden et al.(Hecksteden, 2017)) and dynamically updating biomarker reference ranges (Roshan et al.(Roshan et al., 2021)).

Interestingly, these approaches all have a clear endpoint of interest (e.g., cancer diagnosis,(McIntosh et al., 2024; McIntosh et al., 2025) positive doping test, (Sottas et al., 2007) fatigued status,(Hecksteden, 2017), etc. In several applied sport science settings, monitoring of the athlete on a weekly or bi-weekly basis is frequently conducted as part of the training regimen without a clear outcome.(McLean et al., 2010) For example, McLean et al.(McLean et al., 2010) identified that rugby athletes still exhibit suppressed neuromuscular output 48-hours post-match which is useful information for the strength and conditioning practitioners, as changes to the training program can be made to mitigate further decrement. However, this information is not clearly mapped to a specific outcome and rather serves as observational information to help guide the decisions of coaches and industry professionals. Consequently, analysis that allows the practitioners to account for the uncertainty that the observed test is indeed outside of the normal range provides the opportunity to have conversations with coaches about what interventions might make sense given larger or smaller levels of confidence.

Both frequentist and Bayesian approaches seek to determine whether a change - which requires some adjustment from the coach/practitioner - has occurred.(Hecksteden, 2017; Schechter & Adler, 1988) The underlying assumptions and the process followed to achieve those goals are where we see the difference. The relative merits of the approaches have been debated ad nauseum(Bland & Altman, 1998), and so we will not revisit it in this paper. Instead, we want to provide a quick background for adopting a Bayesian approach. As mentioned in the introduction, the inference we need to make is the likelihood that a change has occurred, given the score we have today.(Gelman et al., 1995)

The philosophical difference here is important. With the Bayesian approach, we estimate the likelihood of a change and provide this information to the coach, allowing them to factor in contextual knowledge.(Nayak & Kundu, 2001) For example, while there may be a reasonable statistical likelihood of a change, the coach might adjust this based on prior knowledge about the athlete. Conversely, the coach could also amplify that likelihood based on other contextual factors. This differs from some other approaches, which rely on purely statistical methods to flag changes. To be clear, we are not suggesting that a probabilistic approach cannot be applied with other metrics. However, we believe that the Bayesian philosophy aligns closely with how practitioners think & make decisions. It also ensures that all available information is maintained through to the person who ends up making the decision.

Ultimately, a likelihood-based approach can provide a critical advantage by supporting practitioners as they incorporate their own knowledge into the overall decision-making processes.(Nayak & Kundu, 2001) Whether identifying a specific endpoint (e.g., cancer diagnosis(McIntosh et al., 2024; McIntosh et al., 2025) or doping violation(Sottas et al., 2007)) or guiding weekly training regimens,(MacLehose & Hamra, 2014) these Bayesian methods ensure that all relevant information reaches the decision-maker, allowing more calibrated and informed decisions.

## Practical Example

Longitudinal testing is conducted in applied sport settings as a means of gathering insights about an athlete’s current physiological state.(Heishman et al., 2020) As with all testing, both systematic and random error are elements of the measurement protocol.(Altman & Royston, 2006; Nayak & Kundu, 2001) When observing weekly measurements in athletes, such error clouds the primary question that practitioners are attempting to answer, *“If this week’s observation is substantially different from last week’s observation, how certain can we be that this change is real and requires an intervention?”* Consequently, an individual’s *True Score* on a test can be summarized as a combination of the score observed on the test plus error (Table 1).(Altman & Bland, 1983) One approach to account for error and incorporating prior knowledge about the test procedure is to use Bayes Theorem.(Bland & Altman, 1998; MacLehose & Hamra, 2014; Schechter & Adler, 1988) Briefly, Bayes Theorem offers an analytics approach to combining prior information about the measurement, both the population center and error in the test, and combines that information with the athlete’s observation to calculate a posterior value that can be used to produce a posterior distribution of potential True Values for the athlete. (Gelman et al., 1995)

While Bayes Theorem applied to continuous outcomes - which make up many of the serial measurements in sport - are often mathematically complex and computationally intensive, simple algorithms can be leveraged if the practitioner is willing to make some underlying assumptions regarding the distribution of the data (e.g., normality and homoscedasticity).(Schechter & Adler, 1988) These simple algorithms are not only easy to implement in spreadsheet software but also provide quick results that allow practitioners to make decisions about the estimated true value for an athlete. Additionally, the posterior distribution affords the practitioner the opportunity to make probabilistic statements regarding the observed changes.(Gelman et al., 1995) The aim of this practical example is to provide the practitioner with a simple method of evaluating repeated testing in athletes while accounting for both systematic and random test error.

### Data Simulation

To demonstrate the use of Bayesian updating in a sport science setting we simulate weekly testing of peak concentric power for an athlete who is tested in the countermovement jump twice a week.(Heishman et al., 2020) The athlete is asked to perform three countermovement jumps with arm swing. The average peak concentric power of the three jumps is used for analysis. To make the simulation more *real world* we’ve added missing values for two of the testing periods. The parameters used to create the simulation are available in **Table 2**. A plot of the simulated testing data is seen in **Figure 1**.

**Table 2.** Model Parameters for Simulation

| Variables | Values |
| --- | --- |
| Number of Tests | 45 |
| Average Peak Power | 5000 |
| Weekly Trend | 2 |
| Seasonl Variation | 100 |
| Random Noise | 150 |

**Figure 1.**
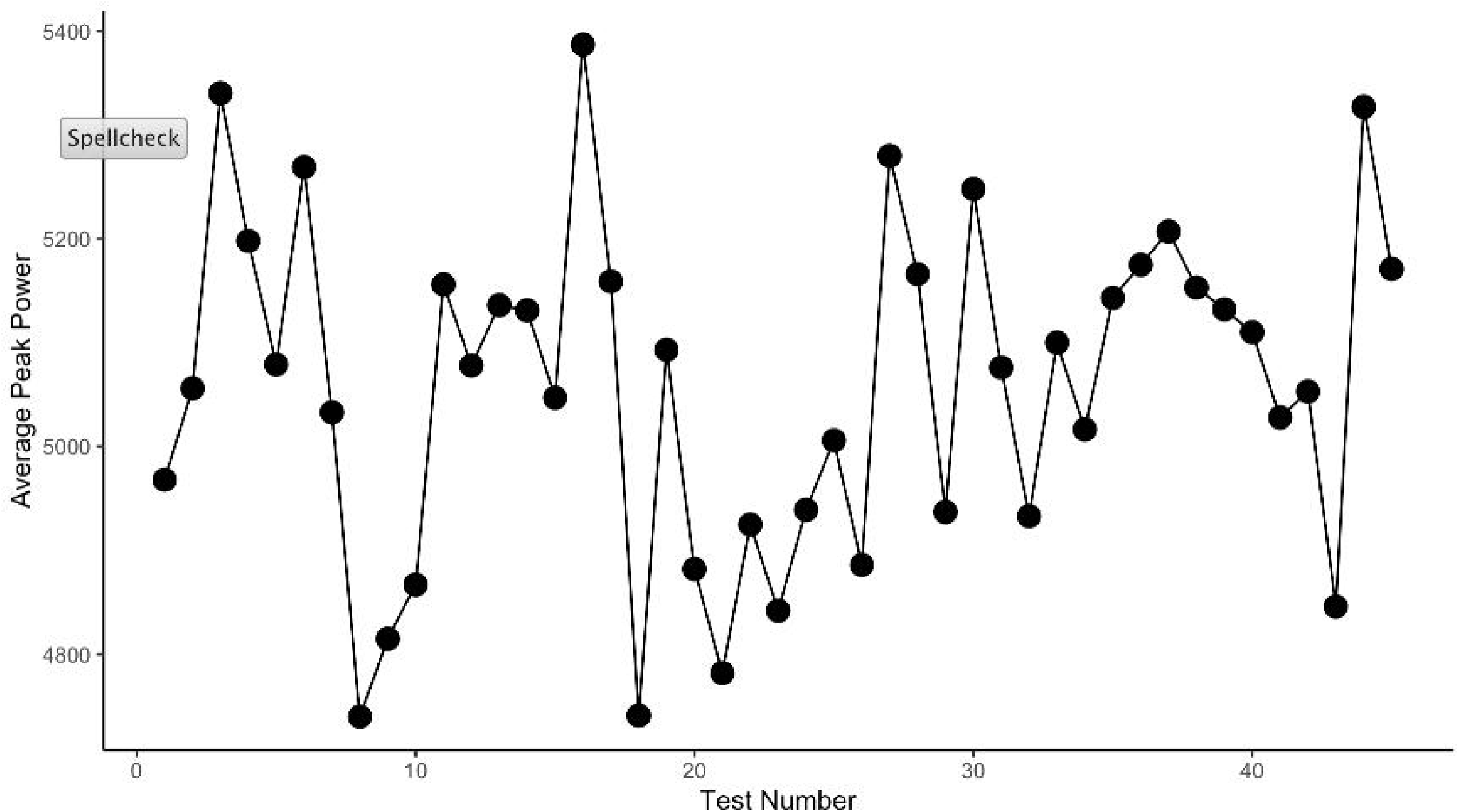
Serial Measurement of Peak Power.

The practitioner’s goal is to identify changes in weekly test scores that are outside of normal – i.e. larger than a smallest meaningful change - to plan interventions or suggest changes to weekly training that may help the athlete recover and perform at a higher level. Unfortunately, this goal is complicated by the fact that the true change observed on a given day is the aggregation of the athletes true score plus some amount of error. A plot of the test-to-test differences and the pooled standard deviation between subsequent tests can be observed in **Figure 2**.

**Figure 2.**
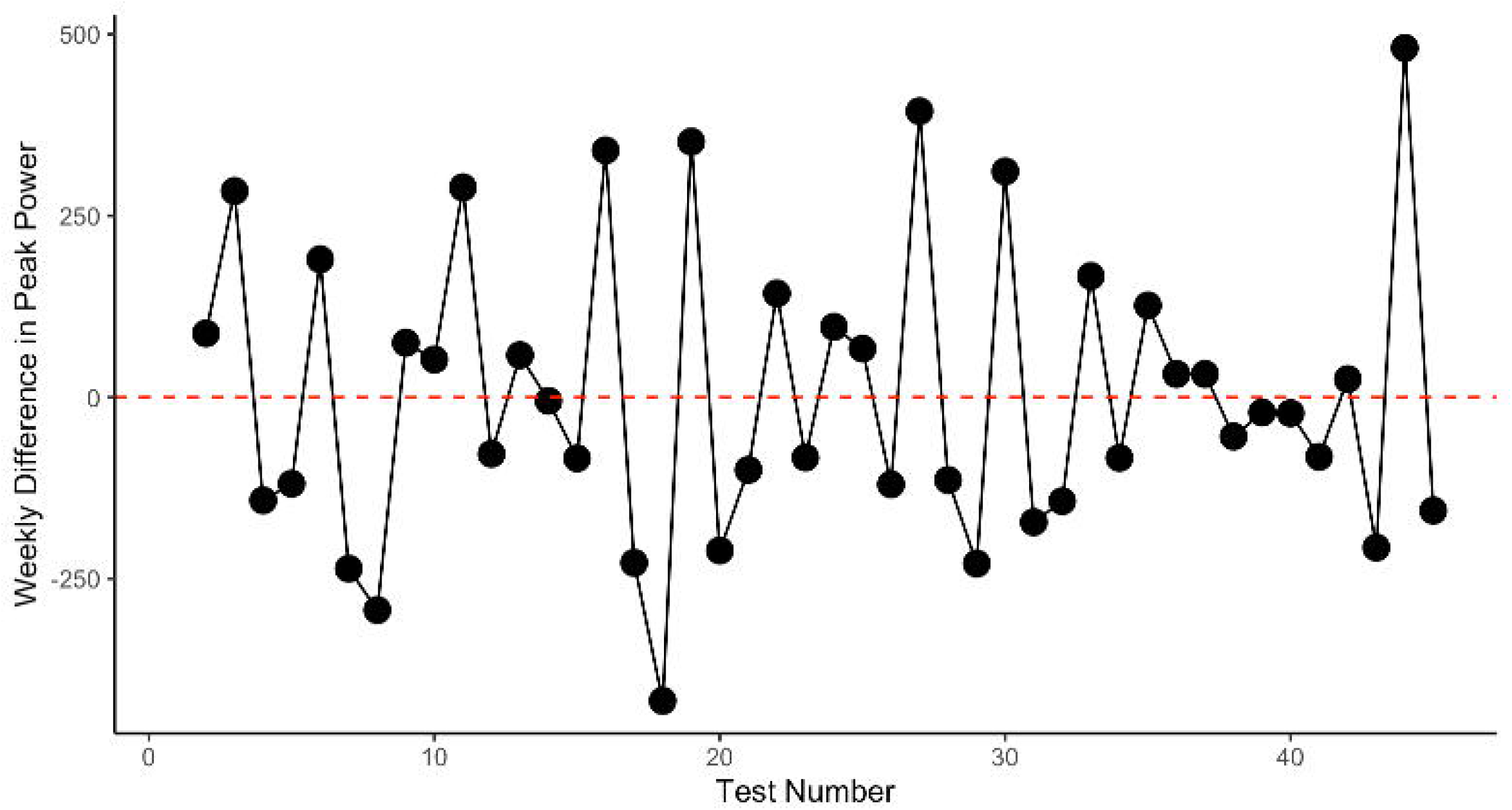
Test-to-test change plot.

To add greater context to the visual, we plot the observed peak concentric power for each test along with the rolling 5-test average and rolling 5-test standard deviation (**Figure 3**). The rolling 5-test average and standard deviation were selected as a pragmatic balance for this situation - long enough to reflect meaningful physiological adaptation (2–3 weeks), but short enough to remain sensitive to acute changes when testing 2x’s a week.

**Figure 3.**
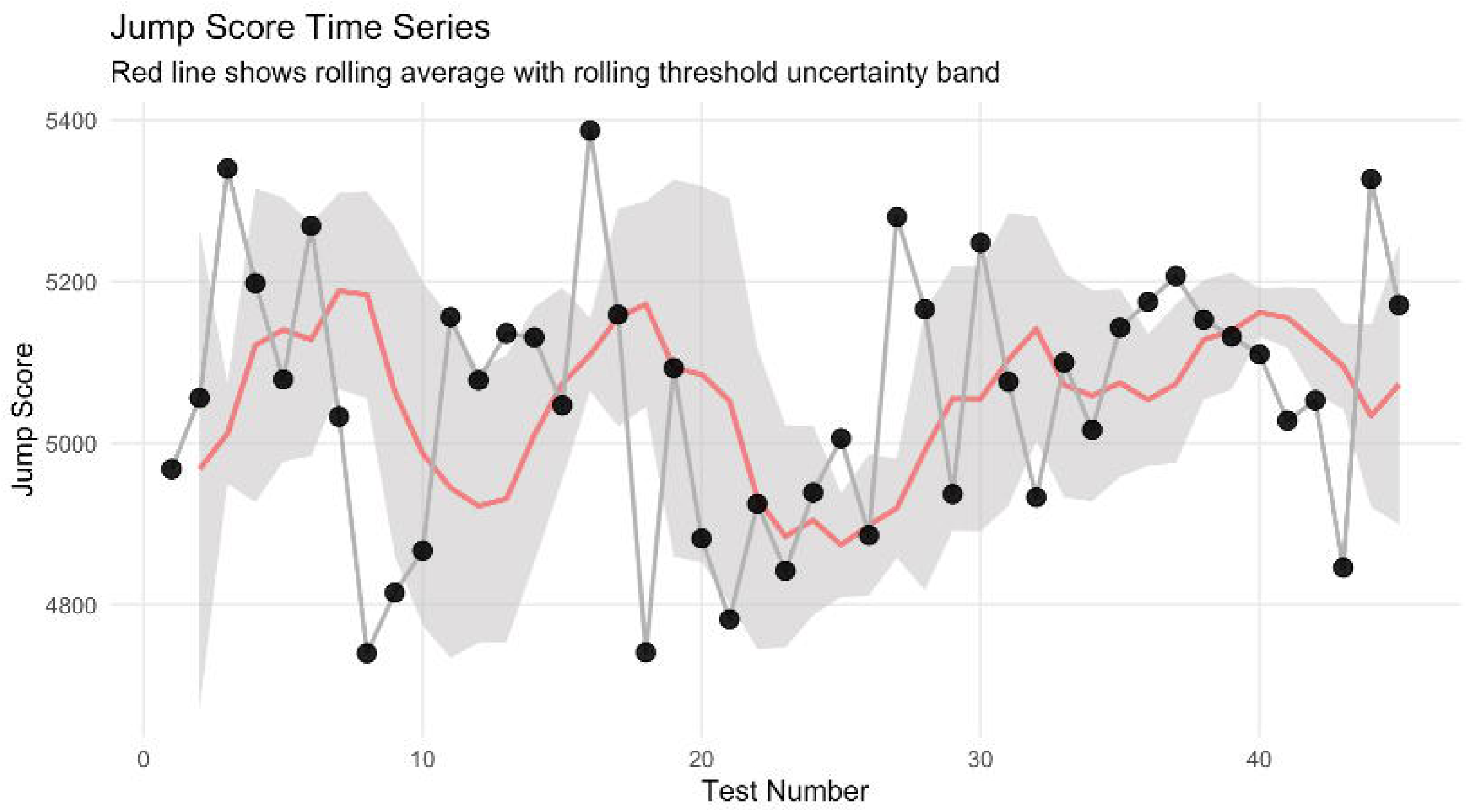
Repeated Countermovement Jump Testing of An Individual Athlete Over Time with Rolling Average. The red line depicts the rolling average with the grey shaded area reporting the uncertainty. Blue points with numerical probability represent points above and red points with probability represent below the Smallest Worthwhile Change for Peak Concentric Power from Heishman et al. (2020).

### Obtaining Standard Error of Measurement and Minimal Detectable Change

The most important aspect of Bayesian analysis is the information contained in the prior.(Gelman et al., 1995) The prior is used to weight the evidence of the observed score to obtain a more reasonable estimate of the true value, termed the posterior. Priors can be selected using subjective expert judgement, pilot studies, or prior research.(Gelman et al., 1995) In this example, we obtain the Smallest Worthwhile Change and Intra-class Correlation Coefficient of Peak Concentric Power reported by Heishman and colleagues who explored the intra-day reliability of several Countermovement Jump variables in NCAA basketball players (**Table 3**).(Heishman et al., 2020) Since our interest is in making an inference about the observed change in subsequent tests, we will specify our prior for the mean change to be 0. That is to say, our prior belief is that athletes exhibit little to no change from one test to the next.

**Table 3.** Prior parameters for Peak Concentric Power from Heishman et al (2018).

| **Variables** | **Values** |
| --- | --- |
| Number of Subject | 22 |
| Number of Measurements | 44 |
| Typical Error | 99.2 |
| Intra-Class Correlation Coefficient | 0.834 |
| Smallest Worthwhile Change | 296.2 |

**Table 4.**
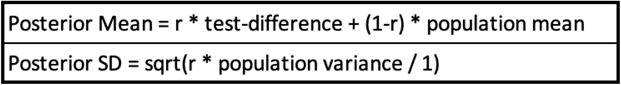
Posterior mean and standard deviation using observed data and prior parameters. Where r is the prior ICC, test-difference is the difference between two tests, population mean is the prior for the mean population test-retest difference (in this case we set it to 0), and population variance is the prior for Typical Error.

|  |
| --- |
| Posterior Mean = $r * \text{test-difference} + (1-r) * \text{population mean}$ |
| Posterior SD = $\text{sqrt}(r * \text{population variance} / 1)$ |

### Calculating the posterior

There are a number of approaches one can use to combine the observed data with the prior parameters. To make this tutorial palatable for the practitioner we will refrain from writing mathematical notation and instead provide the details of the underlying math in plain English. The interested reader who would like to investigate other potential approaches to estimating the posterior with this type of data and better understand the mathematical proofs are encouraged to consult external resources.(Bolstad & Curran, 2016; Gelman et al., 1995) We combine the observed data and prior parameters using the approach taken by Schechter(Schechter & Adler, 1988) to obtain a posterior mean and standard error or measure.

### Drawing Inference

After calculating the posterior mean and standard error of measure for each test difference we plot the results to observe how the prior parameters help to weigh the observed values and pull more extreme values closer to the mean (**Figure 4**). The plot reflects the observed changes against a fixed smallest worthwhile change threshold (grey shaded region), which is the value obtained from Heishman and colleagues.

**Figure 4.**
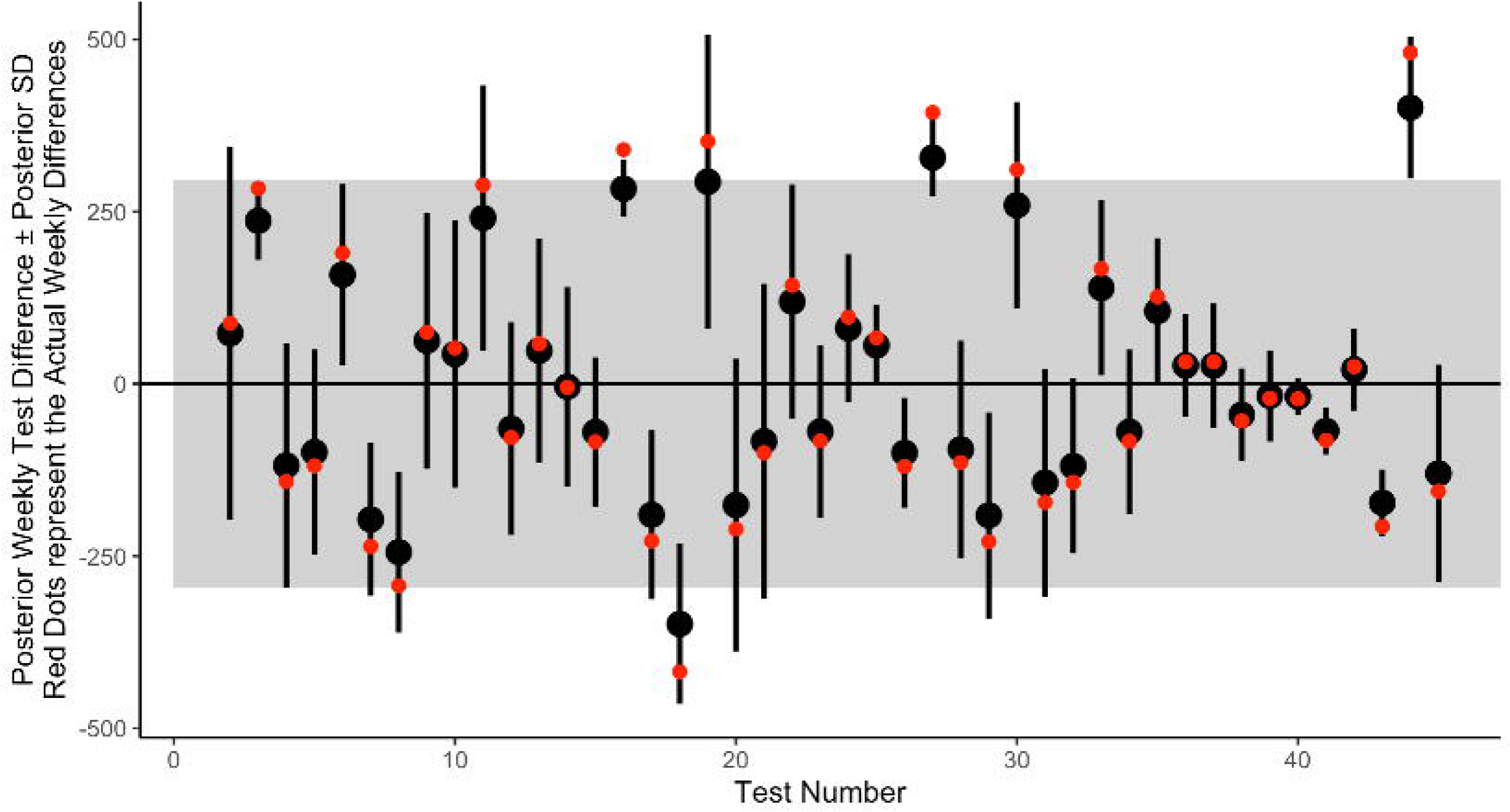
Test-to-test changes in Peak Concentric Power. Red dots represent the observed changes. Block dots represent the Bayesian posterior estimates with error bars representing the posterior standard deviation. The grey shaded region represents the Smallest Worthwhile Change for Peak Concentric Power from Heishman et al. (2020).

Exploring **Figure 4** it is clear that many points are within the normal range of the smallest worthwhile change indicating that a meaningful enough difference from one test to the next has not occurred for the practitioner to be concerned about. However, this plot may be challenging for practitioners given the data is not reflected in the actual units of measurement, but rather differences from test-to-test. Such a plot prevents a clear observation of trends over time. To aid the practitioner, we recommend a time series plot, as seen in **Figure 3**, with the added context of coloring the points when a meaningful change from one test to the next has been observed. Here, we create a simulation for every observation using the Bayesian posterior mean and posterior standard deviation and provide the probability of each change being greater than or less than a threshold of interest, defined as the 5-test rolling standard deviation. The probability of a positive or negative change exceeds 50%, we flag that point in blue or red, respectively, so that the practitioner can further investigate and gather more information about the athlete’s status. An example of such a plot can be seen in **Figure 5**.

**Figure 5.**
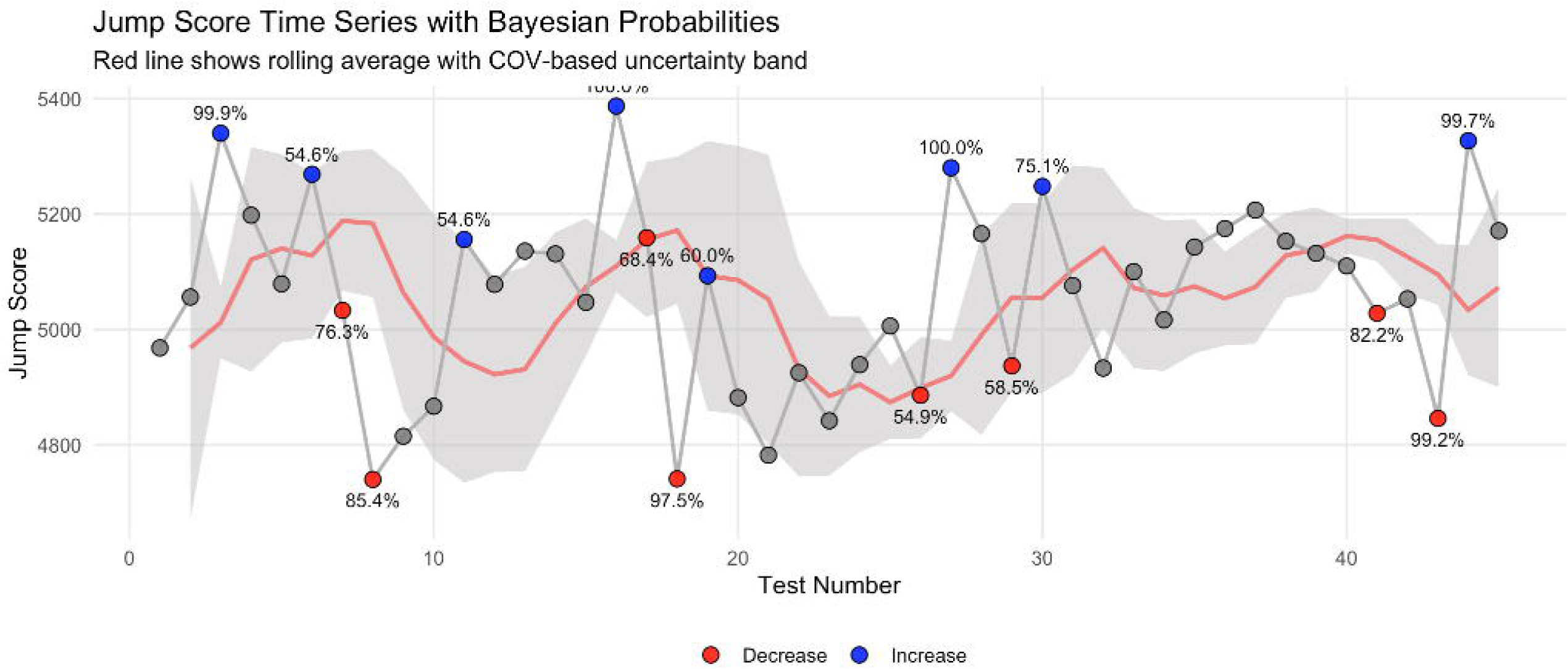
Time Series Plot with Bayesian Posterior Simulation to Determine the Probability That a Change is Greater Than (Blue) or Less Than (Red) the Meaningful Threshold (Rolling 5-Test Standard Deviation).>

Individual observations can be further emphasized by visualizing a histogram of a Monte Carlo Simulation using the posterior mean and standard deviation and the smallest worthwhile change as threshold lines (Appendix). For example, in **Figure 6** we see a full posterior distribution for observation 20. Here, the probability that the change in peak concentric power from test 19 to test 20 was below the threshold of interest (rolling 5-test standard deviation of 233) was 38.3%. The visual is easy for the practitioner to understand as the shaded region reflects the density of the distribution (38.3%) that resides below the threshold of interest.

**Figure 6.**
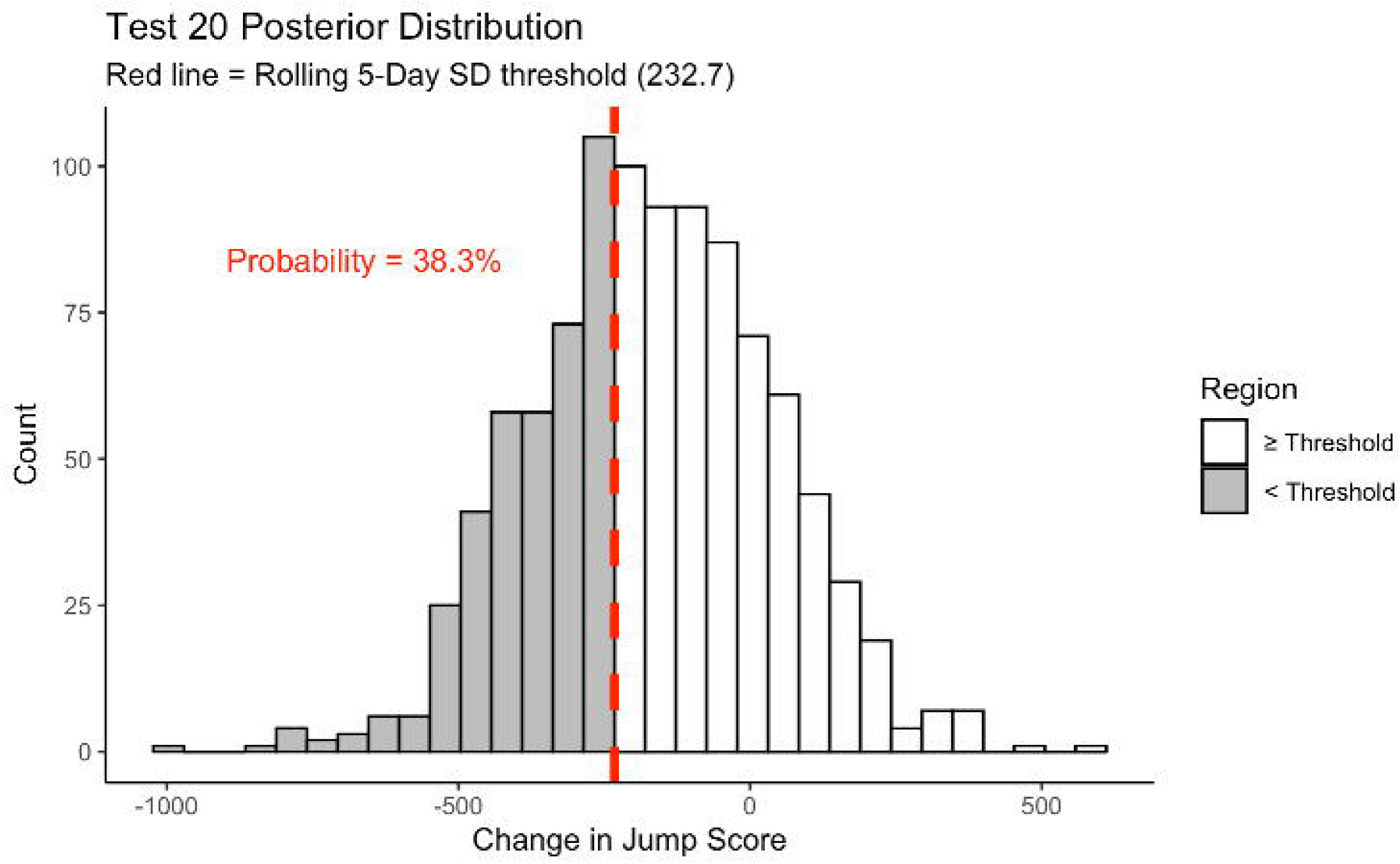
Bayesian Posterior Distribution of Change in Counter Movement Jump. The red dashed line represents the probability of a true changed occurred.

Finally, we can also create team level evaluations per testing jump for each athlete (**Figure 7**), allowing for easy to identify meaningful changes for every athlete within the squad. This information prevents the practitioner from thinking about changes in a binary way and instead facilitates more meaningful discussions about when to intervene in a player’s(West et al., 2025) training based on the probability that a true difference has been observed. This decision may be impacted by multiple factors that weren’t included in the monitoring such as the player’s status on the team (starter vs back up) or time of the season (early season versus playoffs), where some risks might be more acceptable than others.(West et al., 2025) Ultimately, this approach ensures that the decision maker has the full posterior information from the monitoring tests while acknowledging that there are other contextual factors which should influence the final decision. These approaches are designed to ensure that all available information is delivered to the decision maker so that they can make a contextually informed decision. This contrasts with the common criticism that sports scientists make significant decisions based solely on narrowly defined criteria, thereby ignoring critical contextual elements.

**Figure 7.** Changes in Counter Movement Jump Rolling Average Per Individual Athlete by Team. The red dashed lines depict the smallest worthwhile change. The blue points with numerical probability represent points above and red points with probability represent below the individual athlete’s threshold of interested based on the rolling 5-test standard deviation up to but not including this most recent test during the season.

## Practical Applications

Bayesian estimates of change offer an easy and efficient way for data driven exercise and sports scientist practitioners to evaluate the likelihood that a true change has occurred during repeated testing measures. These analyses can be used as an early warning system or to confirm whether a positive or negative change truly occurred based on an observed measurement. This paper provides specific examples from the author’s experience in using Bayesian change point analysis in the sport field and adjoining simulated data and code to reproduce these results, and more importantly, use as a template for the practitioner’s own exercise and sport data.

## Conclusion

Sports scientists have a difficult job. One of the hardest is evaluating individual athletes over time and making good decisions in a high stake’s environment. Many approaches taken to evaluate serial measurements dichotomize data before it gets to the decision maker,(Altman & Royston, 2006) which leads to decisions that failed to consider the probability of all potential outcomes. In contrast, the simple individual Bayesian method demonstrated here provides practitioners with the full posterior distribution, improving information for training, practice, and competition decisions. This comprehensive & probabilistic approach better informs knowledge users tasked with data driven decisions in sport.

## Data Availability

All data produced in the present work are contained in the manuscript.

## Acknowledgements

None

## Open Science

All statistical code is adjoined to as a supplement to this manuscript.

## References

1. Altman, D. G., & Bland, J. M. (1983). Measurement in medicine: the analysis of method comparison studies. Journal of the Royal Statistical Society Series D: The Statistician, 32(3), 307–317.

2. Altman, D. G., & Royston, P. (2006). The cost of dichotomising continuous variables. Bmj, 332(7549), 1080.

3. Armada-Cortés, E., Benítez-Muñoz, J. A., San Juan, A. F., & Sánchez-Sánchez, J. (2022). Evaluation of neuromuscular fatigue in a repeat sprint ability, countermovement jump and hamstring test in elite female soccer players. International journal of environmental research and public health, 19(22), 15069.

4. Beltz, A. M., Wright, A. G., Sprague, B. N., & Molenaar, P. C. (2016). Bridging the nomothetic and idiographic approaches to the analysis of clinical data. Assessment, 23(4), 447–458.

5. Bland, J. M., & Altman, D. G. (1998). Bayesians and frequentists. Bmj, 317(7166), 1151–1160.

6. Bolstad, W. M., & Curran, J. M. (2016). Introduction to Bayesian statistics. John Wiley & Sons.

7. Brown, C. E. (1998). Coefficient of variation. In Applied multivariate statistics in geohydrology and related sciences (pp. 155–157). Springer.

8. Bullock, G. S., Ward, P., Hughes, T., Thigpen, C. A., Cook, C. E., & Shanley, E. (2023). Using Randomized Controlled Trials in the Sports Medicine and Performance Environment: Is It Time to Reconsider and Think Outside the Methodological Box? journal of orthopaedic & sports physical therapy, 53(6), 331–334.

9. Datson, N., Lolli, L., Drust, B., Atkinson, G., Weston, M., & Gregson, W. (2022). Inter-methodological quantification of the target change for performance test outcomes relevant to elite female soccer players. Science and Medicine in Football, 6(2), 248–261.

10. Gelman, A., Carlin, J. B., Stern, H. S., & Rubin, D. B. (1995). Bayesian data analysis. Chapman and Hall/CRC.

11. Harry, J. R., Hurwitz, J., Agnew, C., & Bishop, C. (2024). Statistical tests for sports science practitioners: identifying performance gains in individual athletes. The Journal of Strength & Conditioning Research, 38(5), e264–e272.

12. Hecksteden, A. (2017). A new method to individualize monitoring of muscle. ijspp, 2016, 0120.

13. Heishman, A. D., Daub, B. D., Miller, R. M., Freitas, E. D., Frantz, B. A., & Bemben, M. G. (2020). Countermovement jump reliability performed with and without an arm swing in NCAA division 1 intercollegiate basketball players. The Journal of Strength & Conditioning Research, 34(2), 546–558.

14. Kinsella, D., Fell, J., Berto, C., Robertson, S., & Mole, J. (2012). Analyses and comparison of counter-movement jump performance and self-rated recovery in state under-18s Australian Rules Football players during a national championship.

15. MacLehose, R. F., & Hamra, G. B. (2014). Applications of Bayesian methods to epidemiologic research. Current Epidemiology Reports, 1, 103–109.

16. McDonald, S., Vieira, R., & Johnston, D. W. (2020). Analysing N-of-1 observational data in health psychology and behavioural medicine: a 10-step SPSS tutorial for beginners. Health psychology and behavioral medicine, 8(1), 32–54.

17. Mchunu, N. N., Mwambi, H. G., Rizopoulos, D., Reddy, T., & Yende-Zuma, N. (2022). Using joint models to study the association between CD4 count and the risk of death in TB/HIV data. BMC Medical Research Methodology, 22(1), 295.

18. McIntosh, J. G., Jenkins, M., Wood, A., Chondros, P., Campbell, T., Wenkart, E., O’Reilly, C., Dixon, I., Toner, J., & Martinez-Gutierrez, J. (2024). Increasing bowel cancer screening using SMS in general practice: the SMARTscreen cluster randomised trial. British Journal of General Practice, 74(741), e275–e282.

19. McIntosh, J. G., Wood, A., Jenkins, M., Onwuka, S., Chondros, P., Campbell, T., Wenkart, E., O’Reilly, C., Dixon, I., & Toner, J. (2025). Using an SMS to improve bowel cancer screening: the acceptability and feasibility of a multifaceted intervention. Family Practice, 42(1), cmae073.

20. McLean, B. D., Coutts, A. J., Kelly, V., McGuigan, M. R., & Cormack, S. J. (2010). Neuromuscular, endocrine, and perceptual fatigue responses during different length between-match microcycles in professional rugby league players. International journal of sports physiology and performance, 5(3), 367–383.

21. Nayak, T. K., & Kundu, S. (2001). Calculating and describing uncertainty in risk assessment: the Bayesian approach. Human and Ecological Risk Assessment, 7(2), 307–328.

22. Perini, T. A., Oliveira, G. L. d., Ornellas, J. d. S., & Oliveira, F. P. d. (2005). Technical error of measurement in anthropometry. Revista Brasileira de Medicina do Esporte, 11, 81–85.

23. Roshan, D., Ferguson, J., Pedlar, C. R., Simpkin, A., Wyns, W., Sullivan, F., & Newell, J. (2021). A comparison of methods to generate adaptive reference ranges in longitudinal monitoring. PLoS One, 16(2), e0247338.

24. Sanders, G. J., Boos, B., Rhodes, J., Kollock, R. O., Peacock, C. A., & Scheadler, C. M. (2019). Factors associated with minimal changes in countermovement jump performance throughout a competitive division I collegiate basketball season. Journal of sports sciences, 37(19), 2236–2242.

25. Schechter, C. B., & Adler, R. S. (1988). Bayesian analysis of diastolic blood pressure measurement. Medical Decision Making, 8(3), 182–190.

26. Sottas, P.-E., Baume, N., Saudan, C., Schweizer, C., Kamber, M., & Saugy, M. (2007). Bayesian detection of abnormal values in longitudinal biomarkers with an application to T/E ratio. Biostatistics, 8(2), 285–296.

27. Swinton, P. A., Hemingway, B. S., Saunders, B., Gualano, B., & Dolan, E. (2018). A statistical framework to interpret individual response to intervention: paving the way for personalized nutrition and exercise prescription. Frontiers in nutrition, 5, 41.

28. Turner, A., Brazier, J., Bishop, C., Chavda, S., Cree, J., & Read, P. (2015). Data analysis for strength and conditioning coaches: Using excel to analyze reliability, differences, and relationships. Strength & Conditioning Journal, 37(1), 76–83.

29. Van Der Greef, J., Hankemeier, T., & McBurney, R. N. (2006). Metabolomics-based systems biology and personalized medicine: moving towards n= 1 clinical trials? Pharmacogenomics, 7(7), 1087–1094.

30. Ward, P., Coutts, A. J., Pruna, R., & McCall, A. (2018). Putting the “I” back in team. International journal of sports physiology and performance, 13(8), 1107–1111.

31. Weakley, J., Black, G., McLaren, S., Scantlebury, S., Suchomel, T. J., McMahon, E., Watts, D., & Read, D. B. (2024). Testing and profiling athletes: recommendations for test selection, implementation, and maximizing information. Strength & Conditioning Journal, 46(2), 159–179.

32. Weakley, J., Mann, B., Banyard, H., McLaren, S., Scott, T., & Garcia-Ramos, A. (2021). Velocity-based training: From theory to application. Strength & Conditioning Journal, 43(2), 31–49.

33. West, S., Shrier, I., Impellizzeri, F. M., Clubb, J., Ward, P., & Bullock, G. (2025). Training-Load Management Ambiguities and Weak Logic: Creating Potential Consequences in Sport Training and Performance. Int J Sports Physiol Perform, 20(3), 481–484. 10.1123/ijspp.2024-0158

34. Woodworth, R. J., O’Brien-Malone, A., Diamond, M. R., & Schüz, B. (2016). Happy Days: Positive Psychology interventions effects on affect in an N-of-1 trial. International Journal of Clinical and Health Psychology, 16(1), 21–29.

