## Supplementary material for "Did a true change occur? Improving analytical decisions for serial performance testing in sport": R Code

\# Appendix Code

\# Probability a Change has Occurred - Bayes

\#

\#

\# Simple Proof of Concept

\# •	We are testing CMJ data in an athlete as a measure of "readiness" and "fitness"

\# •	We will use “jump_score” to keep it simple

\# •	We test twice a week and retain the mean jump score (average peak concentric power).

#================================================================================================

\# 1. Load Packages

#================================================================================================

library(tidyverse)

library(ggridges)

library(zoo)

\# set standard plot theme

theme_set(theme_bw())

#===============================================================================================

\# 2. Simulate data

#===============================================================================================

\# Set random seed for reproducibility

set.seed(123)

\# Parameters

n_tests <- 45	# number of total tests performed by the athlete

base_score <- 5000	# average jump score

trend <- 2	# slight upward trend per test

seasonal_amp <- 100	# amplitude of seasonal variation

noise_sd <- 150	# standard deviation of random noise

na_prob <- 0.05	# probability of having missing data

\# Simulate data

simulated_scores <- numeric(n_tests)

for(i in 1:n_tests) {

\# Combine:

\# 1. Base score

\# 2. Slight trend

\# 3. Seasonal variation (sine wave)

\# 4. Random noise

simulated_scores[i] <- base_score +

trend * i +

seasonal_amp * sin(2 * pi * i / 12) + # 12-test cycle

rnorm(1, 0, noise_sd)

}

\# Add some NAs (missing values) randomly

na_positions <- sample(1:n_tests, size = round(n_tests * na_prob))

simulated_scores[na_positions] <- NA

\# Round to whole numbers like the original data

simulated_scores <- round(simulated_scores)

\# Get the total number of jumps

n_jumps <- length(simulated_scores)

#================================================================================================

\# 3. Create the data frame and calculate rolling 5 obseration average for jump score

#================================================================================================

\# Create a data frame with test scores

dat <- data.frame(

test_num = 1:n_jumps,

avg_jump_score = simulated_scores

)

\# Handle missing values by taking the avgerage of last 3 tests

dat <- dat %>%

mutate(

avg_jump_score = ifelse(

is.na(avg_jump_score),

rollapply(avg_jump_score, width = 3, FUN = mean, fill = NA, align = "right", na.rm = TRUE),

avg_jump_score)

)

\# Define window size for rolling calculations

window_size <- 5	# Looking back at previous 5 tests

default_cov <- 4.0 # Default COV when insufficient data (3%)

\# Calculate basic rolling statistics and the difference between consecutive tests

dat <- dat %>%

mutate(

\# Get average of the previous 5 tests

rolling_avg = rollapply(

data = avg_jump_score,

width = pmin(test_num, window_size), # Use fewer tests for early observations

FUN = mean,

fill = NA,

align = "right"

),

rolling_avg = lag(rolling_avg), # Shift to exclude current test

\# Get standard deviation of previous 5 tests

rolling_sd = rollapply(

data = avg_jump_score,

width = pmin(test_num, window_size),

FUN = sd,

fill = NA,

align = "right"

),

rolling_sd = lag(rolling_sd), # Shift to exclude current test

\# Calculate difference between consecutive tests

test_diff = avg_jump_score - lag(avg_jump_score)

)

\# Plot 1: Time series with original values

plot1 <- dat %>%

ggplot(aes(x = test_num, y = avg_jump_score)) +

geom_point(size = 4) +

geom_line() +

labs(x = "Test Number",

y = "Average Peak Power") +

theme_classic()

plot1

\# Plot 2: Plot of Test differences

plot2 <- dat %>%

ggplot(aes(x = test_num, y = test_diff)) +

geom_point(size = 4) +

geom_line() +

geom_hline(yintercept = 0,

linetype = "dashed",

color = "red") +

labs(x = "Test Number",

y = "Weekly Difference in Peak Power") +

theme_classic()

plot2

#================================================================================================

\# 4. Bayesian Priors

#================================================================================================

\## set up the priors from Heishman (2018)

population_mu <- 0	# Prior belief is that the test-to-test difference is 0

r <- 0.834	# ICC from paper

fixed_swc <- 296.2 # SWC value from paper -- used when we don't have a SD for an athlete

\# Incorporating the prior information with the observed data

dat <- dat %>%

mutate(

\# if rolling SD is NA use the fixed_swc from the Heishman paper

rolling_sd = ifelse(is.na(rolling_sd), fixed_swc, rolling_sd),

\# The 5 day rolling SD represents our athletes meaningful threshold

thresh = rolling_sd,

\# Calculate posterior using your formula but with COV-based parameters

posterior_mu = r * test_diff + (1 - r) * population_mu,

posterior_sd = sqrt(r * (thresh^2)/1)

)

#================================================================================================

\# 5. Visualize the Bayesian results

#================================================================================================

\## Plot 3 -- time series of observations and rolling mean and SD

plot3 <- dat %>%

ggplot(aes(x = test_num, y = avg_jump_score)) +

\# Add ribbon for uncertainty using COV thresholds

geom_ribbon(aes(ymin = rolling_avg - thresh,

ymax = rolling_avg + thresh),

fill = "grey",

alpha = 0.5) +

\# add a line for the rolling average

geom_line(aes(y = rolling_avg),

color = "red",

linewidth = 1,

alpha = 0.5) +

\# Add a line for the observed jump score

geom_line(linewidth = 0.8, color = "gray70") +

\# Add points for the observed jump score

geom_point(size = 3,

alpha = 0.9,

na.rm = TRUE) +

labs(title = "Jump Score Time Series",

subtitle = "Red line shows rolling average with rolling threshold uncertainty band",

x = "Test Number",

y = "Jump Score") +

theme_minimal() +

theme(panel.grid.minor = element_blank())

plot3

\## Plot 4 -- test-to-test change against fixed SWC

plot4 <- dat %>%

ggplot(aes(x = test_num, y = posterior_mu)) +

geom_rect(aes(ymin = -fixed_swc, ymax = fixed_swc),

xmin = 0,

xmax = Inf,

fill = "light grey",

alpha = 0.3) +

geom_hline(yintercept = 0,

color = "black") +

geom_errorbar(aes(ymin = posterior_mu - posterior_sd,

ymax = posterior_mu + posterior_sd),

width = 0,

linewidth = 1) +

geom_point(size = 4) +

geom_point(size = 2,

color = "red",

aes(y = test_diff)) +

labs(x = "Test Number",

y = "Posterior Weekly Test Difference ± Posterior SD\nRed Dots represent the Actual Weekly Differences") +

theme_classic()

plot4

\## Plot 5

\# Incorporate the Bayesian Posterior Mu and SD by calculating the

\# probability that the difference from one test to the next is meaningfully

\# outside of the threshold

dat <- dat %>%

rowwise() %>%

mutate(prob_increase = mean(rnorm(n = 1000, mean = posterior_mu, sd = posterior_sd) > thresh, na.rm = TRUE),

prob_decrease = mean(rnorm(n = 1000, mean = posterior_mu, sd = posterior_sd) < -thresh, na.rm = TRUE),

change_interpretation = case_when(prob_increase >= 0.5 ~ "Increase",

prob_decrease >= 0.5 ~ "Decrease",

TRUE ~ "Withing Limits"),

plot_label = case_when(change_interpretation == "Increase" ~ prob_increase,

change_interpretation == "Decrease" ~ prob_decrease))

\# plot the time series data with context about the change taking place

plot5 <- dat %>%

ggplot(aes(x = test_num, y = avg_jump_score)) +

\# Add ribbon for uncertainty using COV thresholds

geom_ribbon(aes(ymin = rolling_avg - thresh,

ymax = rolling_avg + thresh),

fill = "grey",

alpha = 0.5) +

\# add a line for the rolling average

geom_line(aes(y = rolling_avg),

color = "red",

linewidth = 1,

alpha = 0.5) +

\# Add a line for the observed jump score

geom_line(linewidth = 0.8, color = "gray70") +

\# Add points for the observed jump score and colors for meaningful changes

geom_point(aes(fill = change_interpretation),

size = 3,

shape = 21,

alpha = 0.9,

na.rm = TRUE) +

\# Add labels for probabilities

geom_text(aes(label = scales::percent(plot_label, accuracy = 0.1),

vjust = ifelse(prob_increase > 0.5, -1, 2)),

size = 3,

na.rm = TRUE) +

scale_fill_manual(values = c("Increase" = "blue", "Within Limits" = "black", "Decrease" = "red")) +

labs(title = "Jump Score Time Series with Bayesian Probabilities",

subtitle = "Red line shows rolling average with COV-based uncertainty band",

x = "Test Number",

y = "Jump Score",

fill = NULL) +

theme_minimal() +

theme(panel.grid.minor = element_blank()) +

theme(legend.position = "bottom")

plot5

#================================================================================================

\# 6. Analyze a Specific Test Against the Threshold

#================================================================================================

\# Pick a test number to analyze (using test 20 as an example)

obs_test <- dat %>%

filter(test_num == 20)

\# Probability calculation using COV-based threshold

obs_prob <- pnorm(q = -obs_test$thresh,

mean = obs_test$posterior_mu,

sd = obs_test$posterior_sd,

lower.tail = TRUE)

\# Print the probability

cat("Probability below threshold: ", round(obs_prob * 100, 1), "%\n")

\# Simulation

set.seed(7788)

obs_sim <- rnorm(n = 1000,

mean = obs_test$posterior_mu,

sd = obs_test$posterior_sd)

\# Convert to data frame for ggplot

sim_df <- data.frame(value = obs_sim)

\# Calculate probability of being below COV threshold

prob_below_threshold <- mean(obs_sim < -obs_test$thresh)

\# Create the ggplot

obs_plot <- ggplot(sim_df, aes(x = value)) +

\# Add histogram with different colors for regions above/below threshold

geom_histogram(aes(fill = value < -obs_test$thresh),

binwidth = diff(range(obs_sim))/30,

color = "black",

boundary = -obs_test$thresh) +

\# Set colors and legend

scale_fill_manual(values = c("white", "grey"),

name = "Region",

labels = c("≥ Threshold", "< Threshold")) +

\# Add vertical line at threshold

geom_vline(xintercept = -obs_test$thresh,

color = "red",

linetype = "dashed",

linewidth = 1.5) +

\# Add probability text annotation

annotate("text",

x = min(obs_sim) + diff(range(obs_sim))*0.2,

y = max(table(cut(obs_sim, 30)))*0.8,

label = paste0("Probability = ", round(prob_below_threshold * 100, 1), "%"),

color = "red",

size = 4) +

\# Add labels

labs(title = paste0("Test ", obs_test$test_num, " Posterior Distribution"),

subtitle = paste0("Red line = Rolling 5-Day SD threshold (",

round(obs_test$thresh, 1), ")"),

x = "Change in Jump Score",

y = "Count") +

theme_classic() +

theme(legend.position = "right")

\# Display the plot

obs_plot

#================================================================================================

\# 7. Multiple Athletes Comparison with Enhanced COV Visualization

#================================================================================================

\# Set parameters for all athletes

window_size <- 5	# Number of previous tests to use for rolling calculations

default_cov <- 3.0	# Default COV value when insufficient data (3%)

r <- 0.834	# ICC value from Heishman (2018)

#---------------------------------------------------------------------------------------------

\# Step 1: Create function to generate and analyze data for one athlete

#---------------------------------------------------------------------------------------------

generate_athlete_data <- function(athlete_id, base_score = 5000) {

\# Set parameters for simulating this athlete's data

n_tests <- 45	# Number of tests per athlete

trend <- 4	# Overall trend in scores

seasonal_amp <- 400	# Seasonal variation amplitude

noise_sd <- 400	# Random noise standard deviation

\# Generate scores with consistent randomization

set.seed(athlete_id)	# Use athlete_id as seed for reproducibility

simulated_scores <- numeric(n_tests)

\# Create time series with trend, seasonality and noise

for(i in 1:n_tests) {

simulated_scores[i] <- base_score +

trend * i +	# Add trend

seasonal_amp * sin(2 * pi * i / 12) +	# Add seasonality

rnorm(1, 0, noise_sd)	# Add random noise

}

\# Round scores to whole numbers

simulated_scores <- round(simulated_scores)

\# Create initial dataframe for this athlete

dat <- data.frame(

athlete_id = paste("Athlete", athlete_id),

test_num = 1:n_tests,

avg_jump_score = simulated_scores

)

\# Handle missing values and calculate rolling statistics

dat <- dat %>%

\# Handle any missing values

mutate(

avg_jump_score = ifelse(

is.na(avg_jump_score),

rollapply(avg_jump_score, width = 3, FUN = mean, fill = NA, align = "right", na.rm = TRUE),

avg_jump_score)

) %>%

\# Calculate rolling average

mutate(

\# Get average of previous tests (not including current test)

rolling_avg = rollapply(

data = avg_jump_score,

width = pmin(test_num, window_size),

FUN = mean,

fill = NA,

align = "right"

),

rolling_avg = lag(rolling_avg),

\# Calculate test difference

test_diff = avg_jump_score - lag(avg_jump_score),

\# Calculate rolling standard deviation

rolling_sd = rollapply(

data = avg_jump_score,

width = pmin(test_num, window_size),

FUN = sd,

fill = NA,

align = "right"

),

rolling_sd = lag(rolling_sd)

) %>%

\# Calculate COV and threshold

mutate(

\# Calculate COV (Coefficient of Variation)

rolling_cov = ifelse(

is.na(rolling_avg) | is.na(rolling_sd) | rolling_avg == 0 | test_num < 2,

default_cov, # Use default when not enough data

(rolling_sd / rolling_avg) * 100

),

\# Calculate the threshold based on COV

cov_threshold = rolling_avg * (rolling_cov / 100)

)

\# Get only the last test for each athlete for the summary plot

last_obs <- dat %>%

filter(test_num == max(test_num)) %>%

\# Calculate the change from average

mutate(

change = avg_jump_score - rolling_avg,

\# Store raw COV percentage for reference

cov_percentage = rolling_cov

) %>%

\# Calculate posterior using ICC-based approach

mutate(

\# Calculate posterior using ICC (r)

posterior_mu = r * change + (1 - r) * 0, # 0 is population mean (no change)

posterior_sd = sqrt(r * (cov_threshold^2)/1)

)

\# Calculate probabilities using simulation

n_sims <- 1000

set.seed(athlete_id * 100) # Ensure reproducible results

sims <- rnorm(n_sims, mean = last_obs$posterior_mu, sd = last_obs$posterior_sd)

\# Calculate probability of meaningful increase/decrease

last_obs$prob_increase <- mean(sims > last_obs$cov_threshold)

last_obs$prob_decrease <- mean(sims < -last_obs$cov_threshold)

return(last_obs)

}

#---------------------------------------------------------------------------------------------

\# Step 2: Generate data for multiple athletes

#---------------------------------------------------------------------------------------------

\# Generate data for 10 athletes

athlete_data <- map_dfr(1:10, ~generate_athlete_data(.x))

#---------------------------------------------------------------------------------------------

\# Step 3: Create enhanced visualization to compare all athletes

#---------------------------------------------------------------------------------------------

\# Add color and label columns

athlete_plot_data <- athlete_data %>%

mutate(

\# Determine color based on direction of change

point_color = case_when(

prob_increase > 0.5 ~ "Increase",

prob_decrease > 0.5 ~ "Decrease",

TRUE ~ "No Change"

),

\# Create percentage labels for significant changes

prob_label = case_when(

prob_increase > 0.5 ~ sprintf("%d%%", round(prob_increase * 100)),

prob_decrease > 0.5 ~ sprintf("%d%%", round(prob_decrease * 100)),

TRUE ~ ""

),

\# Calculate ratio of change to COV threshold for visualization

change_to_threshold_ratio = abs(change) / cov_threshold,

\# Create label showing COV percentage for each athlete

cov_label = sprintf("COV: %.1f%%", cov_percentage)

) %>%

\# Sort by change magnitude for better visualization

arrange(desc(abs(change)))

#---------------------------------------------------------------------------------------------

\# Step 4: Create visualization with ratio to COV

#---------------------------------------------------------------------------------------------

\# Create a standardized plot showing change as percentage of COV threshold

ratio_plot <- ggplot(athlete_plot_data, aes(y = athlete_id)) +

\# Add zone representing 100% of COV threshold

annotate("rect",

xmin = -1,

xmax = 1,

ymin = 0.5,

ymax = nrow(athlete_plot_data) + 0.5,

fill = "gray90",

alpha = 0.5) +

\# Add reference line at zero

geom_vline(xintercept = 0, color = "black") +

\# Add standard threshold lines at ±100% of COV

geom_vline(xintercept = -1,

color = "red",

linetype = "dashed") +

geom_vline(xintercept = 1,

color = "red",

linetype = "dashed") +

\# Add error bars standardized by COV threshold

geom_errorbarh(aes(xmin = (change - posterior_sd) / cov_threshold,

xmax = (change + posterior_sd) / cov_threshold,

y = athlete_id),

height = 0.2) +

\# Add the actual data points

geom_point(aes(x = change / cov_threshold,

color = point_color),

size = 3) +

\# Add probability labels

geom_text(aes(x = change / cov_threshold,

label = prob_label),

hjust = ifelse(athlete_plot_data$change > 0, -0.3, 1.3),

vjust = -0.8,

size = 3) +

\# Add change values directly next to each point

geom_text(aes(x = change / cov_threshold,

label = sprintf("(Δ=%d)", round(change))),

hjust = ifelse(athlete_plot_data$change > 0, -0.2, 1.2),

vjust = 1.5, # Centered vertically with the point

size = 3,

color = "darkgray") +

\# Configure colors

scale_color_manual(values = c(

"Increase" = "blue",

"Decrease" = "red",

"No Change" = "black"

)) +

\# Add labels and styling

labs(title = "Standardized Changes Relative to COV Threshold",

subtitle = "Values show change as a proportion of each athlete's individual COV threshold",

x = "Change as Proportion of COV Threshold",

y = "Athlete ID",

color = "Change Direction") +

scale_x_continuous(

breaks = c(-2, -1.5, -1, -0.5, 0, 0.5, 1, 1.5, 2),

labels = c("-200%", "-150%", "-100%", "-50%", "0%", "50%", "100%", "150%", "200%")

) +

theme_classic() +

theme(

legend.position = "bottom",

axis.text.y = element_text(size = 10)

) +

\# Use scale_y_discrete to control the order

scale_y_discrete(limits = rev(athlete_plot_data$athlete_id))

\# Display the standardized plot

ratio_plot
